# The Comparison of enhanced recovery after surgery versus traditional pathway in early-onset scoliosis surgery

**DOI:** 10.1101/2023.04.08.23288328

**Authors:** Keyi Jian, Jian Cui, Chunbin Li, Rong Liu

## Abstract

The optimized enhanced recovery after surgery (ERAS) pathway in patients with early onset scoliosis (EOS) has not yet been comprehensively described. This study explored the efficacy and feasibility of an integral process of the ERAS pathway in posterior spinal fusion (PSF) surgery in EOS patients. A total of 70 patients were included in this study, with 35 patients receiving treatment using an ERAS pathway designed and implemented by a multidisciplinary team. The remaining 35 patients followed the traditional pathway (TP) perioperative care. Patient demographics, radiographic parameters, surgical data, and clinical information were collected and analyzed retrospectively. There were no significant differences in sex, height, weight, age, body mass index, preoperative hemoglobin level, fusion segments, number of screws, Cobb angle of the main curve, or surgical duration between the ERAS and TP groups. Regarding pain intensity, the estimated blood loss (EBL), first ambulation time, length of analgesic use, postoperative length of stay (LOS), drainage duration, drainage volume, and incidence of blood transfusion were significantly lower in the ERAS group than in the TP group. The ERAS pathway in EOS orthopedic surgery effectively reduces intraoperative bleeding, alleviates postoperative pain, reduces complications, accelerates recovery, and shortens hospital stays. Therefore, spinal surgeons should adopt the ERAS pathway in EOS surgery.

## 1. Introduction

The Scoliosis Research Society defines early onset scoliosis (EOS) as all types of scoliosis that develop within 10 years of age[1-2]. Early onset scoliosis, a common congenital disorder that develops in infants and early childhood, manifests primarily as scoliosis and spinal deformity and can lead to severe physical and psychological impairment, affecting the patient’s quality of life[3-4]. According to previous studies, early onset scoliosis is 4–8 cases per 10,000 births, with congenital scoliosis being the most common pediatric spinal deformity[5]. Clinically, common treatments for EOS include surgery, traction, and posterior spinal fusion (PSF) surgery[6]. PSF is a safe and effective procedure for treating early-onset scoliosis. Despite the safety and feasibility of growing fusion rods for EOS, the surgical treatment causes significant pain, trauma, and psychological stress in pediatric patients[7-8]. In addition, a series of complications after surgery, such as bleeding, infection, and nerve function impairment, can lead to prolonged recovery time [9-10]. The challenges of postoperative care for PSF surgery are reducing patient pain and complications from analgesic medications and facilitating recovery from physical trauma.

Kehlet first proposed enhanced recovery after surgery (ERAS) in 1997 as a multidisciplinary and multi-modal method to improve the perioperative prognosis and rehabilitation of inpatients, enhanced recovery after surgery (ERAS) was first proposed by Kehlet in 1997[11-12]. ERAS is a comprehensive system that combines medical, therapeutic, and psychological approaches to accelerate postoperative patient recovery [13].

Previous studies have shown that different specialties have demonstrated the advantages of ERAS in improving postoperative patient recovery and reducing the length of stay (LOS)[14-15]. The ERAS protocol consists of a series of evidence-based perioperative care approaches, such as reducing surgical stress, providing early nutrition, and preventing nausea and vomiting[16]. However, the optimal postoperative recovery approach for patients with early-onset scoliosis has not been systematically or comprehensively described. We compared the effectiveness of the traditional and ERAS pathways in postoperative rehabilitation care for patients with early-onset scoliosis.

## 2. Materials and Methods

Our team received approval from the Ethics Committee of Chengdu No.11 People’s prior to the start of this study. Informed consent was obtained from all study participants. The inclusion criteria were as follows: (1) Patients with EOS undergoing growing rod treatment, (2) those in good physical and psychological condition, (3) those who underwent posterior spinal fusion (PSF) surgery at the end of the growing rod treatment, (4) without vertebral column resection (VCR) osteotomy, and (5) no other neurological disease. The indications for PSF surgery were scoliosis > 50°, Risser sign > level II, onset less than 2 years, and no other disc pathology. The exclusion criteria were haematological disorders, abnormal preoperative hemoglobin levels, neurological abnormalities, poor compliance, and other conditions that prevent following the ERAS pathway.

Our institution began exploring and implementing the ERAS pathway protocol in 2018. The patients received traditional perioperative care (traditional pathway group, TP) before the ERAS protocol was implemented, after which they experienced enhanced recovery after surgery (ERAS group). It should be noted that patients in both groups received the same surgical approach from the same surgical team. Posterior spinal fusion (PSF) surgery after the rod-lengthening procedure is described as the creation of the location of the growing rod and pedicle screws on the dorsal surface and the placement of the screws and growing rod according to a standard surgical path. A vertebrectomy was performed to correct the curves. The treatment of postoperative complications and discharge criteria for all patients were the same.

Before the ERAS protocol was developed and implemented, routine pathways for perioperative care after the same team performed PSF surgery. A multidisciplinary team of specialists, including spine surgeons, nurses, anesthesiologists, psychologists, and nutritionists, designed and implemented a standard ERAS approach based on enhanced recovery after surgery and evidence-based medicine. The protocol was finalized after months of discussion and modification by a multidisciplinary team. A nutritionist and psychologist optimized the patient’s condition by providing assessments of mental health and nutritional status. A comparison of the procedures performed between the two groups is presented in Table 1. The discharge indicators for both groups were similar, including good vital signs and mental status, normal feeding and defecation, no fever or wound infection, pain relief, no interference with daily life, and the completion of rehabilitation training alone.

### 2.1. Outcome measures

We retrospectively collected clinical information, demographic profiles, imaging parameters, and surgery-related data from patients in the ERAS and traditional pathways. Clinical information included pre- and postoperative hemoglobin levels, postoperative pain score (visual analog scale [VAS]), drainage volume, duration of drainage, duration of analgesia, first ambulation time, and length of stay. The demographic information included age, sex, height, weight, and body mass index (BMI). Surgical information included operative time, intraoperative bleeding, fixed segments, and the number of screws. The imaging parameters in our study included the main Cobb angle correction rates before and after surgery. We also analyzed patients’ hospitalizations and postoperative complications.

### 2.2. Statistical analysis

All the statistical analyses were performed using SPSS version 21.0 (IBM Corp., Armonk, NY, USA). Differences in continuous variables and parametric data between the two cohorts were assessed using two independent t-tests and Fisher’s exact test for categorical differences in outcome variables. A p-value of ≤0.05 was considered statistically significant.

## 3. Results

### 3.1. Demographic characteristics

Seventy patients with EOS who underwent PSF surgery were retrospectively reviewed, with 35 in each ERAS and TP group. There were 12 and 13 male patients in the ERAS and TP groups, respectively. ERAS and TP groups had an overall mean age of 13.17±1.44 and 13.86±1.50 years, respectively. The patient demographic characteristics are presented in Table 2. There were no significant differences between the ERAS and TP groups regarding sex, height, weight, age, BMI, or preoperative hemoglobin levels.

### 3.2. Surgical characteristics

Both groups underwent PSF surgery and achieved prominent spinal deformity correction. The main curve Cobb angle was corrected from 89.03±12.51°to 46.74±7.41°in the ERAS group and from 84.86±10.53° to 44.69±4.66°in the TP group. The correction rate in both groups was approximately 50%. The ERAS and TP groups used similar screws and surgical segments, and the difference in the operative time was not statistically significant. However, the ERAS group was significantly less likely to have estimated blood loss than the TP group. Detailed information is provided in Table 3.

### 3.3. Postoperative recovery characteristics

The pain assessment (VAS) was 3.94±0.94 and 2.00±0.64 on postoperative day 1 and day 3 in the ERAS group, both significantly lower than in the TP group (P<0.001). Postoperative hemoglobin was significantly lower in the TP group 107.31±6.68 than in the ERAS group 114.63±6.71 (p<0.001). The drainage volume and duration of drainage in the ERAS group were 209.31±10.42 ml and 2.14±0.60 days, respectively, less than those in the TP group (P<0.001). There was a statistical difference in the duration of analgesic drug application, 2.43±0.78 days in the ERAS group and 4.37±0.81 days in the TP group (p<0.001). The postoperative length of hospital stay was significantly less in the ERAS group than in the TP group (4.66±0.84 vs. 5.97±0.86 days). The time to first ambulation was shorter in the ERAS group at 2.29±0.57 days compared to 4.94±0.73 days in the TP group. Two patients in the ERAS and TP groups underwent allogeneic transfusion, and there was no significant difference between the two groups.

In the TP group, two patients developed wound infection, five developed nausea and vomiting, three developed fever, and eight developed anemia. In contrast, among the 35 patients in the ERAS group, postoperative complications included one case of wound infection, two cases of nausea and vomiting, and one case of fever. The incidence of postoperative anemia was significantly lower in the ERAS group than the TP group. The postoperative recovery characteristics are listed in Table 4.

## 4. Discussion

The ERAS perioperative care protocol was first introduced by the Danish surgeon Henrik Kehle[17]. This program is a concept in which multiple disciplines and specialties work together to improve preoperative surgical tolerance, reduce intraoperative stress, and promote rapid postoperative recovery[18]. Although this concept was initially used only in colorectal surgery, it has now been validated in the perioperative surgical period in various studies [19-21]. Early-onset scoliosis is a common congenital deformity that causes abnormalities in the patient’s appearance, limits thoracic development, affects cardiopulmonary function, and impairs physical and mental health[22-23]. PSF surgery is invasive, with long operating times, risk of neurological complications, and bleeding. In previous studies showing a rapid return to normalcy in patients after scoliosis surgery using the ERAS pathway versus the traditional pathway, the results showed a significant reduction in operative time and bleeding in the ERAS group[24-25]. Gornitzky et al.[26] found a significant decrease in pain scores on POD 0, 1, and 2 and a decrease in opioid analgesic use on POD 0 through the ERAS pathway. Wainwright et al.[27] determined the rationale and modalities for implementing the ERAS pathway in major spinal surgeries through a retrospective study. Wang et al. [28] implemented the ERAS pathway in patients undergoing endoscopic lumbar interbody fusion and observed decreased LOS and blood loss after surgery. During follow-up, patients receiving ERAS had better pain scores and reduced use of pain medications. In previous studies, the main focus was on the conceptual components of ERAS without a comprehensive and refined overall protocol based on the ERAS pathway; therefore, we explored the effectiveness and feasibility of an optimized ERAS pathway in patients undergoing PSF surgery.

In a previous study, the authors investigated the perioperative management of patients with adolescent idiopathic scoliosis using the ERAS pathway[29]. The perioperative management of patients with EOS is a crucial aspect of postsurgical recovery because these patients have worse nutritional status, lower BMIs, and lower hemoglobin levels than adolescent idiopathic scoliosis patients. Based on extensive surgical experience and the characteristics of our pediatric patients, we developed a multi-modal, multidisciplinary care plan for EOS surgery with staff, including nurses, orthopedic surgeons, psychologists, nutritionists, and anesthesiologists. Patients were evaluated in detail before surgery, and targeted adjustments and functional training were designed based on the evaluation results to improve patient tolerance to the surgery. Preoperative communication with patients and their families to answer questions about the procedure improved patient compliance with the ERAS pathway and eliminated preoperative anxiety. Patients with scoliosis were often accompanied by psychological anxiety and depression during treatment. Spinal deformity correction helped improve the patient’s psychological condition. Therefore, patient self-confidence should improve before surgery. During the operation, we tried to refine the procedure as much as possible to avoid damaging the microvasculature and other vessels, reduce intraoperative bleeding, and keep the intraoperative body temperature constant. Promoting patient recovery and preventing complications are important. When surgery was completed, patients received satisfactory pain management and rehabilitation training.

Tranexamic acid, or controlled lowering of blood pressure, was used to reduce bleeding during surgery. Tranexamic acid reduces intraoperative bleeding, improves hemoglobin levels, and increases the risk of thrombosis[30]. Our research found that using TXA resulted in significantly less drainage and bleeding and a lower probability of transfusion in the ERAS group than in the TP group, with higher hemoglobin levels.

The ERAS guidelines emphasize postoperative analgesia given to patients through multiple modalities. Postoperative pain management has always been a difficult aspect of spinal surgery. Effective postoperative pain management can reduce a patient’s stress response, promote early mobility to the floor, and accelerate healing and recovery. In the ERAS group, postoperative pain was reduced using a postoperative analgesic device (butorphanol + sufentanil), COX-2 inhibiting analogs, and incisional infiltration anesthesia. Our findings revealed that pain scores on the first and third postoperative days were significantly less in the ERAS group than in the TP group.

The length of stay for postoperative patients through the ERAS pathway is 1-2 days shorter than that through the traditional pathway[31]. Length of stay has been used to indicate the condition of care and the merits of rapid recovery. We found that patients in the ERAS pathway were discharged earlier, facilitating recovery and reducing hospitalization costs. With the ERAS pathway, patients had better preoperative surgical tolerance, treatment compliance, and nutritional status. Postoperative pain management allows patients to move earlier, facilitates wound recovery, and reduces nausea and vomiting. These factors led to shorter hospital stays. Regarding complications, anemia was more likely to occur in the TP group.

Although our study provides valuable insights into the efficacy and feasibility of implementing an ERAS pathway in PSF surgery for EOS patients, some limitations must be acknowledged. Firstly, the sample size of our patients was small. Secondly, the study was retrospective, which may have introduced potential biases. Another limitation is that the same surgical team performed the surgeries, which may have influenced the outcomes. Therefore, future multicenter prospective studies with larger sample sizes must further validate our findings and address these limitations.

## 5. Conclusions

Based on our research, we found that implementing the ERAS pathway in EOS orthopedic surgery effectively reduced intraoperative bleeding, alleviated postoperative pain, reduced complications, accelerated recovery, and shortened hospital stays. Therefore, spinal surgeons should adopt the ERAS pathway in EOS surgery.

## Data Availability

All data produced in the present study are available upon reasonable request to the authors.

