## Supplemental Table for "The Comparison of enhanced recovery after surgery versus traditional pathway in early-onset scoliosis surgery"

**Table 1.** The protocol of ERAS pathway and traditional pathway.

| TP Group | Procedures | ERAS Group |
|---|---|---|
| Pre-operative preparation | | |
| General information about surgery-related conditions | Consultation and Answers | Eliminate patients' preoperative fears and anxieties by informing them about the surgery, including the procedure, the risks and complications associated with the surgery, and answering their confusions |
| Brief introduction of perioperative-related precautions | Education and Communication | Introduce patients to the process and principles of the ERAS pathway, including various preparations for the perioperative period, including diet, pain management, psychological adjustment, and rehabilitation training, as a way to improve compliance with the pathway |
| General assessment: demographic characteristics, such as age, height, weight, etc., vital signs, such as blood pressure, heart rate, oxygen saturation, etc<br>Hematologic assessment: electrolyte balance status, coagulation status<br>Assessment of cardiopulmonary status | Assessment | General assessment: demographic characteristics, such as height, weight, BMI, and age, and vital signs, such as blood pressure, oxygen saturation, and heart rate<br>Hematologic assessment: electrolyte balance status, coagulation status<br>Assessment of cardiopulmonary status<br>Pain intensity assessment<br>Self-nutritional status assessment<br>Self-functional assessment<br>Mental Health Assessment |
| Not applicable | Special training before surgery | Stair climbing for aerobic and functional exercise |

| | | |
|---|---|---|
| | | Balloon blowing training to improve lung function |
| | | Improve spinal mobility by traction and extension of the spine |
| | | Preoperatively by drinking a highly concentrated solution drink, such as 10% glucose solution 5ml/kg |
| Prohibited within 8 hours before the start of induction of anesthesia | Preoperative intestinal preparation | Management of postoperative bloating through the use of pro-gastrointestinal motility drugs |
| | | Removal of solid food within 6 hours prior to the start of induction of anesthesia |
| | | Removal of gastrointestinal water within 2 hours prior to the start of induction of anesthesia |
| Intraoperative procedures | | |
| General positioning | Surgical site positioning | Elastic decompression is applied to the parts of the skin in contact with the surgical platform (knees, ankles, elbows, wrists, chest, anterior superior iliac spine, etc.) to reduce prolonged compression of the body weight on the peripheral nerves |
| | | Posture adjustment of the patient to minimize thoracoabdominal pressure |
| General anesthesia, the exact type of anesthesia depends on the anesthesiologist | Anesthesia mode | The anesthesia induction phase was administered by rocuronium (0.6 mg/kg, iv), propofol (2.5 mg/kg, iv), midazolam (1-2 mg, iv), and sufentanil (0.1-0.5 mg/kg, iv) |

| | | |
|---|---|---|
| | | Anesthesia was sustained by segmental remifentanil (0.2ug/kg/min, iv) and propofol (9-15mg/kg, iv) |
| Intraoperative prevention is not performed | Anti-infection prevention | Antibiotics are given 30 min after the start of the procedure and additional antibiotics are used for procedures with a total duration of more than 3h |
| | | Anesthesia induction 30 min later by opioids such as light ketones, 0.1-0.2 mg/kg and COX-2 inhibitors (e.g., parecoxib, 40 mg, iv) |
| Analgesic drug selection based on physician's personal preference | Pain Management | Dexmedetomidine (0.4 mg/kg/hour, iv), propofol (target-controlled infusion, 4-12 mg/kg/hour) and remifentanil (0.1-0.3 mg/kg/minute, iv) for sustaining phase therapy |
| | | The surgery is carried out delicately to avoid neuromuscular blocking damage. |
| | | Intraoperative cytological salvage |
| | | Monitor arterial pressure in real time and perform controlled hypotension anesthesia when the mean arterial pressure is 70-75 mmHg |
| Blood product transfusions are used to treat patients with anemia such as hemoglobin <70g/L. | Blood Management | TXA (pre-dermal incision soaking dose 20mg/kg + infusion 10mg/kg/h + 3g TXA topical application) |
| | | Blood product transfusions are used to treat patients with anemia such as hemoglobin <70g/L. |

| | | |
|---|---|---|
| No prevention of hypothermia | Temperature management | Maintain a core body temperature of 36 degrees Celsius through a multi-modal approach, such as the use of warm air blowers, airway humidification, warm infusion of fluids, warm blankets, and increased room temperature in the operating room |
| Drug selection based on physician's personal preference | Intraoperative fluid management | Rigorous fluid therapy based on infusion targets |
| Subfascial drainage | Leads Management | Subfascial drainage |
| Postoperative care management | | |
| Fasting for 6 hours after surgery | | Two hours post-op, a small amount of fluid is consumed according to the patient's tolerance and requirements |
| | | Where it is tolerated, a liquid diet is started at 4-6 hours to supplement nutrition and promote gastrointestinal motility. |
| Fluid diet within 24-48 hours depending on tolerance | Ingestion Management | Normal diet is allowed at POD 1 if the patient does not have PONV |
| Normal eating after 48 hours | | High protein diet on the first postoperative day to improve postoperative nutritional status |
| | | Supplementation of iron and folic acid according to the patient's hemoglobin level |
| Metoclopramide (10 mg, intramuscular) when nausea and vomiting occur | Management of postoperative gastrointestinal-related complications | Postoperative dual antiemetic prophylaxis (ondansetron, 4 mg + dexamethasone, 10 mg IV) |

| | | |
|---|---|---|
| | | Metoclopramide (10 mg, intramuscular) when nausea and vomiting occur |
| Removal of catheter after walking on the ground | Postoperative rehabilitation program | Encourage early postoperative activity and ambulation |
| Early activity after removal of drainage tube | | Removal of catheter after recovery from anesthesia |
| | | Remove drainage when drainage is <100ml |
| Postoperative 30 days to move to the ground with brace protection | | Attempt to go to the floor on postoperative day 3-5 |

*COX-2, cyclooxygenase 2; i.v., intravenous injection; TXA, tranexamic acid; b.i.d., twice daily; POD, postoperative day; PONV, postoperative nausea and vomiting; ERAS, enhanced recovery after surgery; TP, traditional pathway.

**Table 2. Demographic characteristics between two groups.**

| | TP group | ERAS group | P value |
|---|---|---|---|
| Sample size | 35 | 35 | - |
| Gender(M: F) | 13:22 | 12:23 | 1.000 |
| Height | 158.51±6.04 | 158.00±6.18 | 0.726 |
| weight | 52.80±4.42 | 52.14±3.05 | 0.471 |
| Age (y) | 13.86±1.50 | 13.17±1.44 | 0.055 |
| BMI (kg/m2 ) | 21.02±1.32 | 20.93±1.42 | 0.797 |
| Preoperative hemoglobin level (g/L) | 115.09±6.10 | 115.63±7.05 | 0.732 |

BMI, body mass index; TP, traditional pathway; ERAS, enhanced recovery after surgery.

**Table 3. Surgical information.**

| | TP group | ERAS group | P value |
|---|---|---|---|
| Fusion segment | 11.09±1.87 | 11.17±1.90 | 0.850 |
| Screw number | 22.06±3.58 | 22.23±3.67 | 0.844 |
| Preoperative cobb of main curve (°) | 84.86±10.53 | 89.03±12.51 | 0.136 |
| Postoperative Cobb of main curve (°) | 44.69±4.66 | 46.74±7.41 | 0.170 |
| Correction rate (%) | 0.47±0.05 | 0.47±0.05 | 0.749 |
| Estimate blood loss (ml) | 401.71±38.08 | 313.29±39.20 | 0.000* |
| Surgical duration (min) | 276.14±24.62 | 274.86±28.50 | 0.841 |

*, P<0.05; TP, traditional pathway; ERAS, enhanced recovery after surgery.

**Table 4. Postoperative recovery characteristics and complications.**

| | TP group | ERAS group | P value |
|---|---|---|---|
| Postoperative recovery characteristics | | | |
| VAS of POD 1 | 4.74±0.85 | 3.94±0.94 | 0.000* |
| VAS of POD 3 | 3.06±0.73 | 2.00±0.64 | 0.000* |
| POD 1 hemoglobin level (g/l) | 107.31±6.68 | 114.63±6.71 | 0.000* |
| Drainage duration (day) | 3.74±0.70 | 2.14±0.60 | 0.000* |
| Drainage volume (ml) | 428.57±109.33 | 209.31±10.42 | 0.000* |
| Analgesic medicine (day) | 4.37±0.81 | 2.43±0.78 | 0.000* |
| Postoperative LOS (day) | 5.97±0.86 | 4.66±0.84 | 0.000* |
| First ambulation time (day) | 4.94±0.73 | 2.29±0.57 | 0.000* |
| Early complications | | | |
| Wound infection | 2 | 1 | 0.986 |
| Nausea and vomiting | 5 | 2 | 0.839 |
| Fever | 3 | 1 | 0.900 |
| Allogeneic blood transfusion | 2 | 2 | 1.000 |
| Anemia | 8 | 0 | 0.060 |

*, $P<0.05$; TP, traditional pathway; ERAS, enhanced recovery after surgery, POD, postoperative day; VAS, visual analog score; LOS, length of stay.
